# Conditional Growth References for Vietnamese Children: Quantile Regression of Growth Distributions and Phenotypes

**DOI:** 10.64898/2026.09.23.26363771

**Authors:** Nhan T. Ho

## Abstract

**Background and objective:** The WHO growth reference judges a child’s height against the population at one age, without using the child’s own earlier growth. This study built and validated a quantile regression based conditional growth reference predicting a child’s future height for age distribution from early growth status.

**Methods:** Using deidentified longitudinal annual school children health check data from three major cities in Vietnam, this study linked a baseline height and BMI for age Z score near age 6 to outcome visits near age 12 or 15 in 4,857 children. A linear quantile regression and a pooled quantile generalized additive model, fit by sex, were compared against a linear benchmark and a naive persistence model on a held out test set, using pinball loss, interval coverage, and width against the WHO 90% band. This study also tested adding growth phenotype and checked generalization across the three cities.

**Results:** The pooled model outperformed the discrete quantile regression in every group. Every individualized 90% interval was narrower than the WHO band, by 34.6 to 54.5%. Growth phenotype improved accuracy in most groups. The model generalized well to a held out Hanoi or Haiphong but poorly to a held out Ho Chi Minh City, where coverage fell to between 0.35 and 0.52, though the all-city model stayed well calibrated for Ho Chi Minh City itself.

**Conclusion:** Conditional quantile modeling gives individualized growth predictions markedly narrower than a fixed population reference and gains further from growth phenotype, but needs local validation before use at an unrepresented site.

## INTRODUCTION

Height-for-age remains one of the most widely used indicators of child health, and the World Health Organization’s growth reference for school-aged children and adolescents is the global benchmark against which an individual child’s height is judged ^1^. That reference is built from the population distribution at each age, so it tells a clinician or a parent where a child stands relative to peers of the same age and sex, but it says nothing about where that specific child is likely headed given their own earlier growth. Two children can share an identical height-for-age Z score at age 6 and still be on genuinely different paths by adolescence, depending on their own prior trajectory, yet a single fixed reference band treats them identically.

Conditional growth analysis was developed to close part of this gap, and Vietnam already has a substantial body of work in this tradition. In the PRECONCEPT cohort mostly in young children before school age, conditional measures of linear and ponderal growth, essentially each child’s own deviation from their expected trajectory, predicted attained height and BMI at school age better than unconditional measures alone, with faster early linear growth most strongly associated with later height for age and conditional weight gain tracking more closely with later BMI for age and overweight risk ^2^. The same conditioning logic extended to non-anthropometric outcomes, linking early linear growth to better intellectual functioning and fewer emotional difficulties in school-age children ^3^. These studies establish that a child’s own growth history carries real, usable predictive information beyond a single cross-sectional measurement. This is the similar premise this paper builds on but instead using early school age growth trajectories to predict pubertal growth outcomes.

A separate methodological tradition, quantile regression, offers a way to go further than conditioning on the mean alone. Rather than estimating a single expected value, quantile regression models the entire conditional distribution of an outcome, letting the association between a predictor and an outcome vary at the low, middle, and high ends of that outcome’s range ^4^. In the region, this approach has mostly been used to study heterogeneous downstream consequences of growth rather than growth itself. Young Lives quantile analyses across Ethiopia, India, Peru, and Vietnam found that mid-childhood linear growth mattered most for human capital outcomes at the lowest deciles of schooling and vocabulary scores ^5^, and a quantile decomposition of Vietnam’s economic boom showed that income growth reached most of the child population but left the most severe nutritional deficits largely unmoved ^6^.

What is missing from this literature is a tool that applies both ideas directly to the growth trajectory itself, an individualized, distributional growth reference rather than a single conditional mean or an analysis of some other downstream outcome. Also there is also a lack of report using early school age growth trajectories to predict pubertal growth outcomes. This gap matters more given how unevenly growth patterns are distributed within Vietnam. Rural infants in Khanh Hoa province fell into lower percentiles of the WHO standards than urban infants under identical criteria ^7^, and weight growth trajectories across the region diverge by country and by rural or urban residence in both timing and magnitude ^8^. A growth reference built without attention to this heterogeneity risks being accurate on average while performing poorly for specific subgroups or sites.

This paper addresses the above gap directly, building on a cohort of school children in three major cities in Vietnam (Hanoi, Hochiminh city, Haiphong) that has already characterized the population pubertal growth spurt, the distributional association between BMI and height-for-age, the distributional association between prepubertal BMI and pubertal growth velocity, and distinct peri-pubertal growth phenotypes in this same cohort ^9–14^. Using quantile regression, this study constructs and validates a conditional growth reference that predicts a Vietnamese child’s future height-for-age distribution, not just its expected value, from their own early height and BMI status, and this study evaluates its calibration, its added value over a fixed population reference, the incremental contribution of growth phenotype, and whether it generalizes across the cohort’s three cities.

## METHODS

### Study population and prior work

This retrospective analysis uses the multi-city longitudinal annual school health check dataset from Hanoi, Ho Chi Minh City, and Haiphong, Vietnam, collected from 2018 to 2025 and described in detail in earlier papers from this program ^9–14^. Height-for-age and BMI-for-age Z scores were calculated using the WHO 2007 growth reference for school-aged children and adolescents ^1^, following the same construction used throughout the series.

This study was approved by Ethical Committee of Vinmec International Hospital (approval number 0231/2024/CN/HDDD VMEC) with a waiver of individual informed consent as the study used de-identified routinely collected retrospective school health check data. The study, methods and report were carried out in accordance with the Declaration of Helsinki and STROBE (STrengthening the Reporting of OBservational studies in Epidemiology) guideline. STROBE checklist with this study corresponding status was included in related file.

### Analytic sample

Eligible children needed one valid anthropometric visit in a conditioning window of age 5 to 9 years, used to define a single baseline height-for-age Z score (HAZ) and BMI-for-age Z score (BAZ) closest to age 6, and at least one valid visit within 1 year of a nominal target age of 12 or 15 years, used as the outcome. Age 18 was considered but dropped after a feasibility check showed the cohort’s calendar span could not bridge a 5 to 9 year baseline to an 18 year outcome for the same child. Eligibility required only one visit within each window rather than a second visit inside a narrow spacing interval, since an earlier stage of this program showed that this kind of narrow-window feature can sharply and non-randomly shrink the analytic sample. Children were split 80 to 20 at the child level into training and held-out test sets, stratified by sex and target age.

### Statistical models

The primary model was a linear conditional quantile regression ^4^, fit separately by sex and target age, of target-age HAZ on baseline HAZ, baseline BAZ, a centered baseline age term, and study city, across a grid of quantiles from 0.05 to 0.95. A second, pooled quantile generalized additive model, fit with the qgam package ^15^ built on the mgcv framework ^16^, was fit separately by sex across all target ages together, using penalized smooth terms for baseline HAZ, baseline BAZ, and the child’s actual outcome age, so predictions could vary continuously with age instead of only at discrete target ages. Four further models were fit for comparison: an ordinary linear regression benchmark using the same predictors as the primary model, a naive persistence model using each child’s own baseline HAZ as the predicted median, a quantile regression adding a baseline HAZ by BAZ interaction term, and a quantile regression using natural cubic splines on baseline HAZ and BAZ. Where available, each child’s peri-pubertal growth phenotype from a companion functional clustering analysis in this program ^10^ was added as an optional predictor to test its incremental value over the baseline model.

### Model evaluation

All models were evaluated on the held-out test set using median absolute error, root mean squared error, pinball loss across the fitted quantile grid, and empirical coverage of the 50, 80, and 90% central prediction intervals against their nominal levels. An approximate continuous ranked probability score was computed as twice the mean pinball loss across the fitted quantile grid ^17^, and a weighted interval score ^18^ was computed for the pooled model, the only one fit at a quantile grid spanning all three interval widths. Cross-validation used 10-fold resampling at the child level along with a leave-one-city-out procedure to check generalization to a city withheld entirely from training. All analyses were conducted in R ^19^, using the quantreg ^20^, qgam ^15^, mgcv ^21^, and arsenal ^22^ packages.

## RESULTS

### Cohort characteristics and attrition

Of the 97,030 children in the source cohort, 74,914 had at least one visit with both height-for-age and BMI-for-age Z scores available, and 39,301 had a usable visit in the age 5 to 9 year conditioning window (**Supplementary Table S1 Panel A**). After linking each child’s conditioning-window visit to an outcome visit at a target age, the final analytic frame held 4,857 children, 2,445 boys and 2,410 girls contributing to the target age 12 submodel, and only 194 boys and 190 girls contributing to the target age 15 submodel (**Supplementary Table S1 Panel B, Supplementary Table S4**). Target age 18 was checked and dropped entirely, since no child in the cohort had both a baseline visit in the conditioning window and an outcome visit near age 18.

The included and excluded children differed in expected ways (**Supplementary Table S3**). Included children had far more visits (median 5, Q1 to Q3 5 to 7, against a median of 2 in excluded children) and much longer follow-up (median 5.1 years against 0.9 years), and were enrolled earlier on average (median first-visit year 2019 against 2020). Boys and girls were about evenly split in the final cohort (2,447 boys and 2,410 girls) (**Table 1**) and did not differ meaningfully by baseline age, city distribution, or the availability of a second conditioning-window visit or a growth phenotype label, of which 98.0% of children had one available. Baseline height for age and BMI for age Z scores were both higher in boys than in girls (median height for age Z 0.418 against 0.264, median BMI for age Z 0.914 against 0.243, both p < 0.001).

**Table 1.** Baseline characteristics of the conditioning-eligible cohort, by sex.

| Characteristic | Male (N = 2,447) | Female (N = 2,410) | Total (N = 4,857) | P value |
| --- | --- | --- | --- | --- |
| City, n (%) |  |  |  | 0.260 |
| Hanoi (HHN) | 1,381 (56.4%) | 1,344 (55.8%) | 2,725 (56.1%) |  |
| Ho Chi Minh City (HCP) | 818 (33.4%) | 848 (35.2%) | 1,666 (34.3%) |  |
| Haiphong (HHP) | 248 (10.1%) | 218 (9.0%) | 466 (9.6%) |  |
| Baseline age, years |  |  |  | 0.146 |
| Median (Q1-Q3) | 6.800 (6.200, 7.400) | 6.900 (6.200, 7.500) | 6.800 (6.200, 7.500) |  |
| Mean (SD) | 6.847 (0.703) | 6.876 (0.702) | 6.861 (0.703) |  |
| Baseline height-for-age Z |  |  |  | < 0.001 |
| Median (Q1-Q3) | 0.418 (-0.297, 1.090) | 0.264 (-0.356, 0.858) | 0.326 (-0.323, 0.999) |  |
| Mean (SD) | 0.408 (0.989) | 0.273 (0.909) | 0.341 (0.952) |  |
| Baseline BMI-for-age Z |  |  |  | < 0.001 |
| Median (Q1-Q3) | 0.914 (-0.311, 2.549) | 0.243 (-0.608, 1.380) | 0.513 (-0.484, 1.940) |  |
| Mean (SD) | 1.190 (1.853) | 0.485 (1.430) | 0.840 (1.694) |  |
| Has a second conditioning-window anchor visit, n (%) |  |  |  | 0.950 |
| No | 932 (38.1%) | 920 (38.2%) | 1,852 (38.1%) |  |
| Yes | 1,515 (61.9%) | 1,490 (61.8%) | 3,005 (61.9%) |  |
| Has a growth phenotype label available, n (%) |  |  |  | 0.979 |
| No | 49 (2.0%) | 48 (2.0%) | 97 (2.0%) |  |
| Yes | 2,398 (98.0%) | 2,362 (98.0%) | 4,760 (98.0%) |  |

Baseline values are measured at the single conditioning-window visit closest to age 6 years, within the 5 to 9 year window. There were no missing values for any characteristic shown. The second-anchor and phenotype rows describe features used opportunistically in the models below and are not eligibility requirements. P values are from the Kruskal-Wallis test for continuous variables and the chi-square test for categorical variables, comparing boys and girls.

City representation shifted sharply between the two target ages (**Supplementary Table S2**). Hanoi supplied 76.4% of the full cohort and still contributed the largest single share of the target age 12 sample, at 56.1%, but contributed no children at all to the target age 15 sample. Ho Chi Minh City, only 18.7% of the full cohort, supplied 89.3% of the target age 15 sample. Target age 15 results should accordingly be read as representative of Ho Chi Minh City and Haiphong children rather than the full three city cohort.

### Model comparison

On the held out test set, the pooled quantile generalized additive model outperformed the discrete linear quantile regression at every sex by target age combination (**Table 2**). Pinball loss fell from 0.154 to 0.141 in boys and 0.133 to 0.120 in girls at target age 12, and from 0.127 to 0.123 in boys and 0.105 to 0.091 in girls at target age 15. Both quantile models clearly beat the naive persistence baseline, which simply carried a child’s own baseline height for age Z forward unchanged, and the ordinary linear benchmark fit for the median only. Adding a height for age by BMI for age interaction term or replacing the linear terms with natural splines gave pinball loss and coverage nearly identical to the simpler additive quantile regression, so neither extra complexity earned its keep at either target age.

**Table 2.** Comparison of prediction models for height-for-age Z score at target ages 12 and 15 years, held-out test set.

| Model | Sex | Target age, y | n (test) | RMSE | MAE | Pinball loss | CRPS | Coverage, 90% |
| --- | --- | --- | --- | --- | --- | --- | --- | --- |
| Naive persistence (predicted = baseline HAZ) | Male | 12 | 486 | 0.655 | 0.502 | 0.161 | 0.321 | 0.928 |
| Naive persistence (predicted = baseline HAZ) | Male | 15 | 38 | 0.582 | 0.445 | 0.144 | 0.288 | 0.842 |
| Naive persistence (predicted = baseline HAZ) | Female | 12 | 485 | 0.596 | 0.470 | 0.147 | 0.294 | 0.911 |
| Naive persistence (predicted = baseline HAZ) | Female | 15 | 30 | 0.395 | 0.322 | 0.103 | 0.206 | 1.000 |
| Linear benchmark (median only) | Male | 12 | 486 | 0.627 | 0.491 | 0.245 | - | 0.922 |
| Linear benchmark (median only) | Male | 15 | 38 | 0.513 | 0.404 | 0.202 | - | 0.947 |
| Linear benchmark (median only) | Female | 12 | 485 | 0.541 | 0.425 | 0.212 | - | 0.932 |
| Linear benchmark (median only) | Female | 15 | 30 | 0.388 | 0.302 | 0.151 | - | 0.967 |
| Quantile regression, baseline features | Male | 12 | 486 | 0.628 | 0.489 | 0.154 | 0.308 | 0.907 |
| Quantile regression, baseline features | Male | 15 | 38 | 0.515 | 0.405 | 0.127 | 0.255 | 0.921 |
| Quantile regression, baseline features | Female | 12 | 485 | 0.541 | 0.423 | 0.133 | 0.266 | 0.918 |
| Quantile regression, baseline features | Female | 15 | 30 | 0.394 | 0.320 | 0.105 | 0.209 | 0.933 |
| Quantile regression, HAZ x BAZ interaction | Male | 12 | 486 | 0.628 | 0.489 | 0.154 | 0.308 | 0.909 |
| Quantile regression, HAZ x BAZ interaction | Male | 15 | 38 | 0.520 | 0.413 | 0.128 | 0.256 | 0.895 |
| Quantile regression, HAZ x BAZ interaction | Female | 12 | 485 | 0.542 | 0.424 | 0.133 | 0.266 | 0.922 |
| Quantile regression, HAZ x BAZ interaction | Female | 15 | 30 | 0.395 | 0.321 | 0.105 | 0.211 | 0.867 |
| Quantile regression, nonlinear splines | Male | 12 | 486 | 0.625 | 0.486 | 0.153 | 0.306 | 0.907 |
| Quantile regression, nonlinear splines | Male | 15 | 38 | 0.519 | 0.413 | 0.132 | 0.264 | 0.842 |
| Quantile regression, nonlinear splines | Female | 12 | 485 | 0.537 | 0.418 | 0.131 | 0.263 | 0.922 |
| Quantile regression, nonlinear splines | Female | 15 | 30 | 0.395 | 0.326 | 0.098 | 0.197 | 0.900 |
| Quantile regression, baseline only (phenotype subset) | Male | 12 | 475 | 0.633 | 0.493 | 0.155 | 0.310 | 0.903 |
| Quantile regression, baseline only (phenotype subset) | Male | 15 | 38 | 0.515 | 0.405 | 0.129 | 0.258 | 0.921 |
| Quantile regression, baseline only (phenotype subset) | Female | 12 | 480 | 0.542 | 0.425 | 0.133 | 0.266 | 0.915 |
| Quantile regression, baseline only (phenotype subset) | Female | 15 | 30 | 0.394 | 0.320 | 0.105 | 0.209 | 0.933 |
| Quantile regression, phenotype enriched | Male | 12 | 475 | 0.526 | 0.405 | 0.124 | 0.248 | 0.891 |
| Quantile regression, phenotype enriched | Male | 15 | 38 | 0.438 | 0.340 | 0.104 | 0.208 | 0.816 |
| Quantile regression, phenotype enriched | Female | 12 | 480 | 0.383 | 0.281 | 0.087 | 0.174 | 0.902 |
| Quantile regression, phenotype enriched | Female | 15 | 30 | 0.409 | 0.348 | 0.101 | 0.201 | 0.900 |
| Quantile GAM, pooled continuous age | Male | 12 | 486 | 0.631 | 0.494 | 0.141 | 0.282 | 0.922 |
| Quantile GAM, pooled continuous age | Male | 15 | 38 | 0.539 | 0.418 | 0.123 | 0.246 | 0.921 |
| Quantile GAM, pooled continuous age | Female | 12 | 485 | 0.532 | 0.412 | 0.120 | 0.239 | 0.930 |
| Quantile GAM, pooled continuous age | Female | 15 | 30 | 0.392 | 0.297 | 0.091 | 0.183 | 0.900 |

Calibration was close to ideal at target age 12 in both sexes (**Figure 2**), with observed coverage tracking the nominal quantile closely across the full grid, and coverage of the 90% interval at 0.907 in boys and 0.918 in girls (**Supplementary Table S5 Panel A**). Calibration was visibly noisier at target age 15, consistent with its much smaller sample rather than a systematic problem. Every individualized 90% interval in the test set was narrower than the fixed 3.29 Z WHO reference band, with a median width of 1.97 Z in boys and 1.73 Z in girls at target age 12, a 34.6 and 45.3% reduction respectively, and narrower still at target age 15 (**Figure 3**, **Supplementary Table S5 Panel B**).

**Figure 1.**
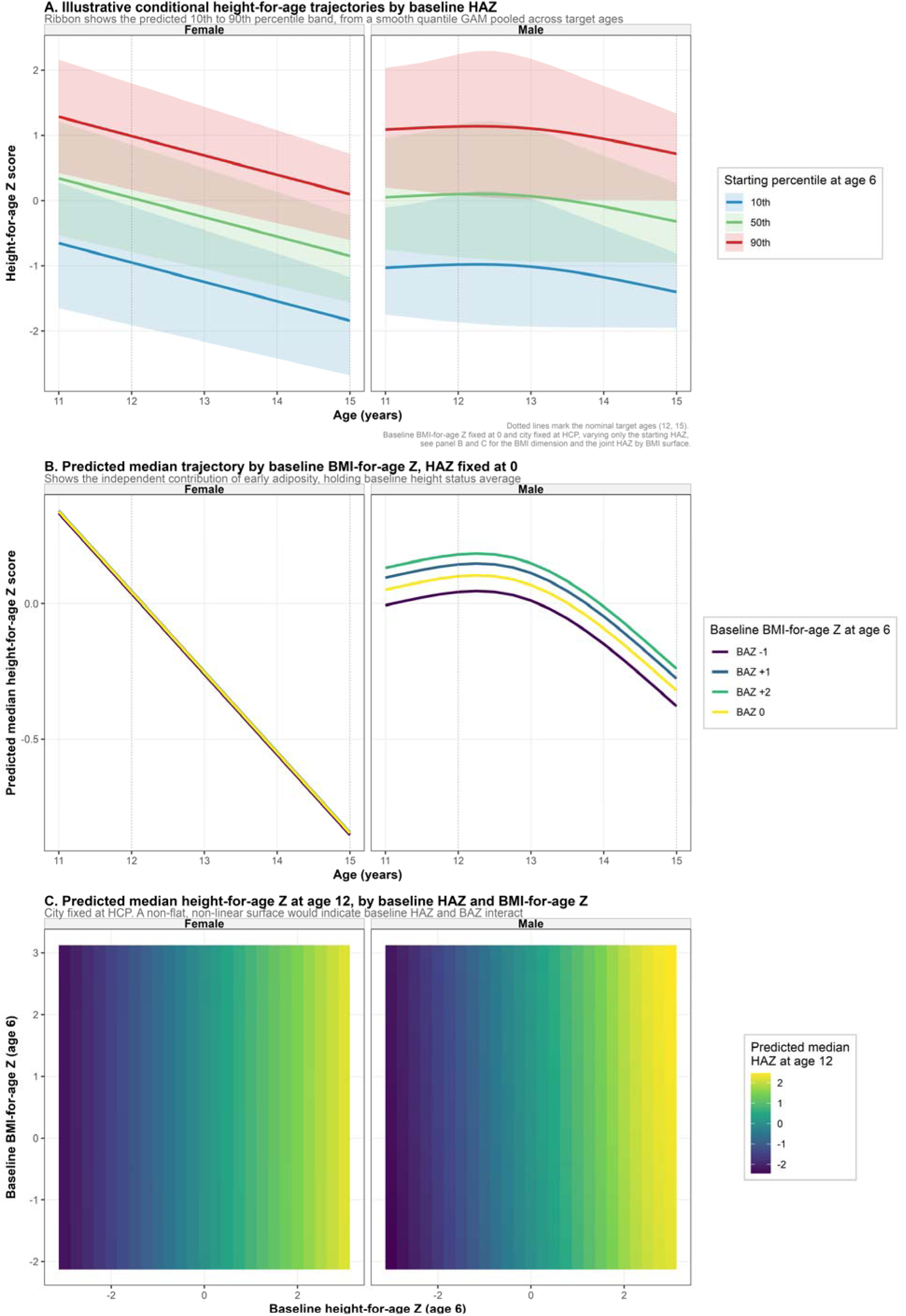
Illustrative conditional height-for-age trajectories from the pooled quantile GAM. Three panels, generated from the same pooled, age-continuous quantile regression model, each varying one baseline predictor at a time and holding the others fixed. **Panel A.** Predicted height-for-age Z trajectory from age 11 to 15 years, for three starting percentiles of baseline height-for-age Z (10th, 50th, and 90th, corresponding to Z of about -1.28, 0, and +1.28 at age 6), faceted by sex (Female, Male). The shaded ribbon shows the predicted 10th to 90th percentile band at each age. Baseline BMI-for-age Z is fixed at 0 and city is fixed at Ho Chi Minh City. Vertical dotted lines mark the nominal target ages, 12 and 15 years, reported in Table 2. **Panel B.** Predicted median trajectory from age 11 to 15 years for four values of baseline BMI-for-age Z (- 1, 0, +1, and +2), with baseline height-for-age Z fixed at 0, faceted by sex. The four lines are visually indistinguishable in the female panel and cleanly, monotonically separated in the male panel. **Panel C.** Predicted median height-for-age Z at target age 12 years, across the joint grid of baseline height-for-age Z (x-axis, approximately -3 to 3) and baseline BMI-for-age Z (y-axis, approximately -2 to 3), faceted by sex, with city fixed at Ho Chi Minh City. Color represents the predicted median height-for-age Z. These panels are illustrative, generated by holding two of the three baseline predictors at fixed reference values rather than showing the full joint predictive distribution for a specific child. Panel C looks flat along the BMI-for-age Z axis, and Panel B confirms why directly: the baseline BMI effect on later median height is essentially absent in girls and present but small, relative to the baseline height effect on the same color scale, in boys.

**Figure 2.**
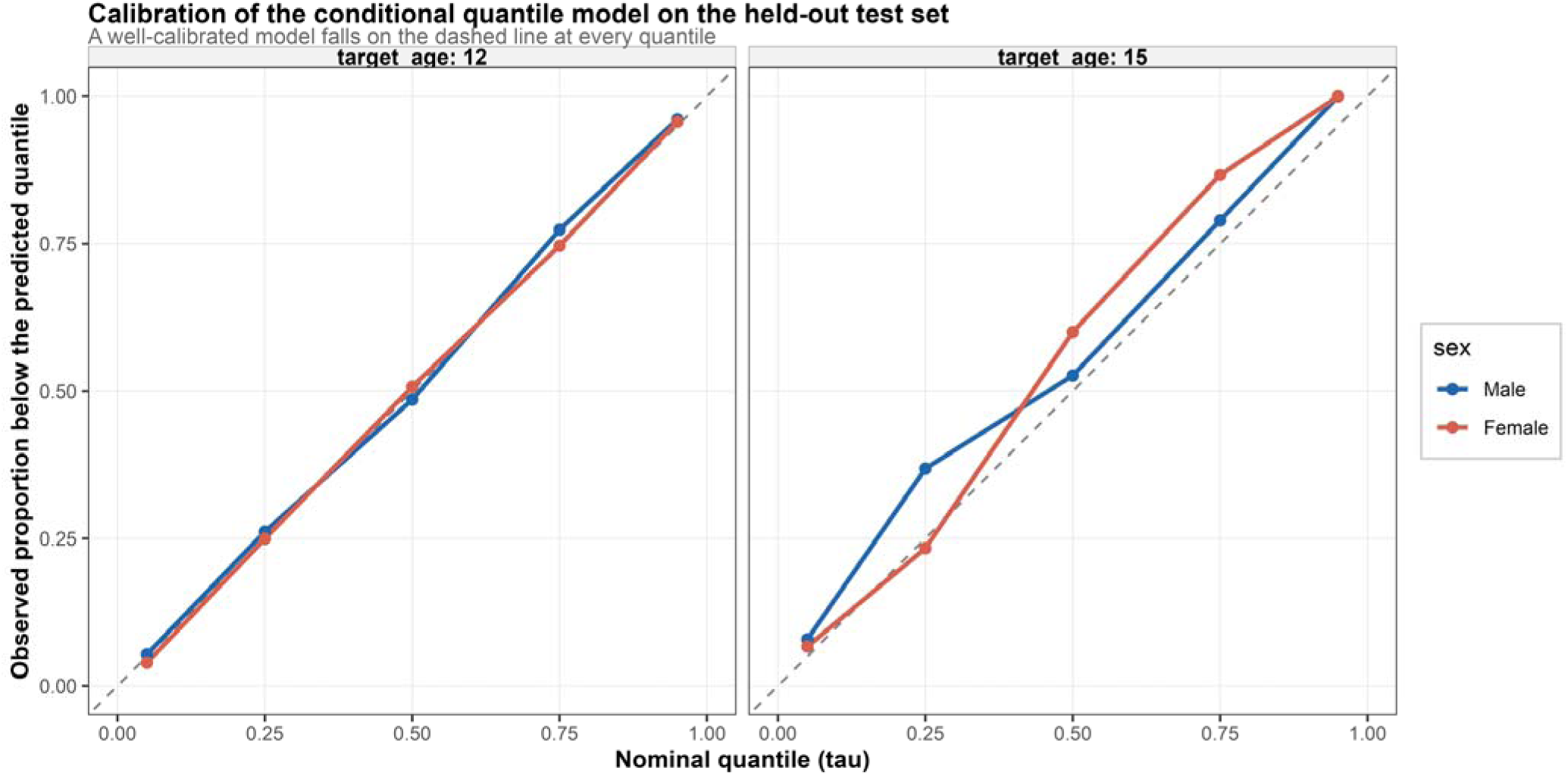
Calibration of the primary quantile regression model on the held-out test set. Nominal quantile (x-axis) plotted against the observed proportion of test-set children whose true height-for-age Z fell below the corresponding predicted quantile (y-axis), separately by sex (Male, Female) and faceted by target age (12, 15). A perfectly calibrated model falls on the dashed diagonal line at every quantile. Calibration closely tracks the diagonal at target age 12, where n is largest (Table 2), and is visibly noisier at target age 15, consistent with its much smaller sample size rather than a systematic modeling problem. Underlying values are in Supplementary Table S5, Panel A, at the 50% and 90% levels, and in S6_calibration_detail at the full fitted quantile grid.

**Figure 3.**
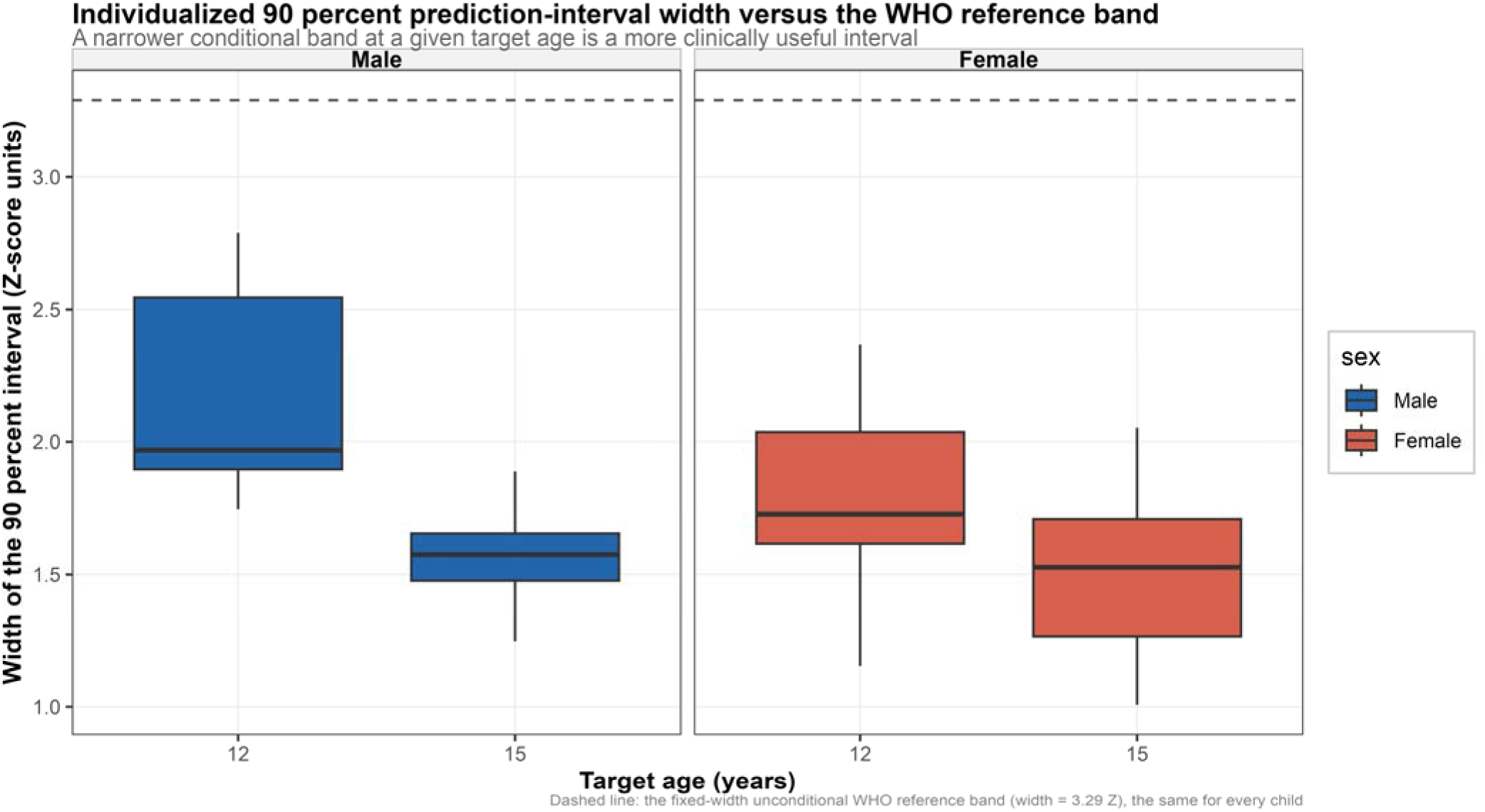
Individualized 90% prediction-interval width compared with the fixed WHO reference band. Boxplots of each test-set child’s own predicted 90% prediction-interval width, in height-for-age Z-score units, by target age (12, 15 on the x-axis) and faceted by sex (Male, Female). The horizontal dashed line marks the fixed width of the unconditional WHO 90% reference band, 3.29 Z, the same for every child regardless of their own baseline status. Numeric summary in Supplementary Table S5, Panel B. Median individualized width ranged from 1.53 to 1.97 Z across the four sex-by-target-age groups, all narrower than the fixed 3.29 Z WHO band, and every single test-set child had an individualized interval narrower than the WHO band (100% at every group).

Adding each child’s growth phenotype label improved median prediction accuracy over the matched baseline model at every combination except girls at target age 15, where the subsample was smallest (**Supplementary Table S6 Panel A**). The largest gain was in girls at target age 12, where mean absolute error fell by 33.8%. A selection effect check found the baseline model performed almost identically on the full test set and on the phenotype available subset alone (**Supplementary Table S6 Panel B**), supporting the phenotype gain as genuine rather than an artifact of an easier to predict subsample.

### Cross validation and site generalizability

Ten fold cross validation gave pinball loss and 90% coverage close to the single test set split results (**Supplementary Table S7 Panel A**). Leave one city out validation showed the model generalized well to a held out Hanoi or Haiphong, but generalized poorly to a held out Ho Chi Minh City, where 90% coverage collapsed to between 0.35 and 0.52 at target age 15 (**Supplementary Table S7 Panel B**). The deployed model, trained on all three cities together, was in fact well calibrated for Ho Chi Minh City specifically, with 90% coverage of 0.91 to 0.96 (**Supplementary Table S7 Panel C**), so this finding is a caution about extending the tool to a genuinely new site rather than a flaw in the model as deployed.

### Illustrative trajectories

The pooled model’s predicted growth trajectories (**Figure 1A**) fanned out sensibly by starting percentile and curved rather than moved in a straight line with age.

Varying baseline BMI for age Z while holding height for age Z fixed (**Figure 1B**) showed four essentially overlapping trajectories in girls and four clearly separated trajectories in boys, and the joint heatmap (**Figure 1C**) showed a corresponding gradient along the height axis with almost none along the BMI axis. Three worked examples illustrate the tool’s individualization directly (**Supplementary Table S8**), a boy who was short for age with a high BMI at age 6 had a predicted 95th percentile of negative 0.92 Z at age 15, showing sustained short stature predicted with high confidence, a distinction a single fixed WHO percentile chart cannot make.

## DISCUSSION

This study built a conditional, quantile-based growth reference for Vietnamese children and found that individualizing on a child’s own early height and BMI status narrowed the predicted range of later height for age well below the fixed WHO band, with phenotype membership adding further accuracy in most groups. These findings sit at the intersection of two literatures that have mostly developed on separate tracks.

The first track, conditional growth analysis, is reported in Vietnam through the PRECONCEPT cohort for young children mainly before school age. Faster linear growth in early life predicts later height for age most strongly, while conditional weight gain tracks more closely with later BMI for age and overweight risk, and each channel carries its own downstream consequences for cognition and mental health ^2,3^. That same channel separation shows up in the present results. Baseline height for age was the dominant predictor of later height for age in both sexes, while baseline BMI for age added only a small, sex specific contribution, significant in boys and essentially absent in girls. Read against the PRECONCEPT findings, this looks less like a modeling quirk and more like a real feature of how linear and ponderal growth propagate differently through childhood, with a growth reference sharing the same conditioning logic already validated in this population.

The second track, quantile regression, has been used in this region mainly to study heterogeneous downstream effects of growth rather than to predict growth itself. The Young Lives quantile analyses found that mid-childhood linear growth mattered most for human capital at the lowest deciles of schooling and vocabulary scores ^5^, and quantile decomposition of Vietnam’s economic boom showed that income gains reached most groups but left the most severe growth deficits largely untouched ^6^. Both point to the same lesson this paper’s calibration and interval width results reinforce directly, an average effect can hide very different pictures at different points in the distribution, and a full conditional distribution is worth reporting rather than a single conditional mean.

A companion literature review of this area found formal quantile regression concentrated in multi-country consortium datasets, with conditional growth analysis common in Vietnam but rarely paired with distributional modeling, and Southeast Asian settings underrepresented outside Vietnam entirely. This paper’s leave-one-city-out result, where a model trained without Ho Chi Minh City data failed badly when applied back to Ho Chi Minh City children, echoes a pattern already documented within Vietnam itself, where rural Khanh Hoa infants fell into lower growth percentiles than urban infants under the same WHO standards ^7^, and where urban and rural weight trajectories diverge in shape across the region more broadly ^8^. A single-country, even single-cohort, growth reference should not be assumed portable across settings without direct validation, and this study’s own data make that point as clearly as the wider literature does.

The short for age, high BMI worked example in this paper, whose predicted interval stayed below zero at age 15 with real confidence, is also consistent with evidence that early growth deficits in term children born small for gestational age tend to persist through mid-childhood rather than resolve on their own, unlike the catch-up typically seen in preterm children born an appropriate size ^23^. A conditional tool that can flag this kind of persistence early, rather than only after it becomes clinically obvious, is exactly the kind of use case this literature points toward.

Vietnam already has some conditional growth research and a smaller quantile regression literature applied to human capital outcomes. This paper connects the two directly to the growth trajectory itself, and its city-level and phenotype findings both point to the same practical implication, that individualized growth monitoring tools built on one region’s data should be validated locally before wider use, and that reporting a full predicted range, not a single expected value, is worth the modest added complexity in a setting like Vietnamese school health programs.

## Data Availability

R codes are available from the corresponding author upon reasonable request. Individual patient-level data cannot be shared due to applicable privacy regulations and the terms of the institutional ethics approval.

## DECLARATION

### Contributor’s statement

Nhan Thi Ho did conceptualization, data curation, formal analysis, investigation, methodology, project administration, resources, software, supervision, validation, visualization, writing original draft, and writing review & editing.

### Conflict of intertest

The author states that there is no conflict of interest.

### Use of Artificial Intelligence

The author performed all original research work regarding scientific content, analyses, interpretations and manuscript writing. The author used AI-assisted tools for language editing and grammar checking during manuscript preparation.

## TABLES

**Supplementary Table S1.** Cohort attrition and the target-age feasibility check.

| ***Panel A. Attrition from the full cohort to the final analytic frame*** |  |  |
| --- | --- | --- |
| **Step** | **n (children)** | **% of full cohort** |
| All children (source cohort) | 97,030 | 100.0 |
| Has a merged height-for-age and BMI-for-age visit | 74,914 | 77.2 |
| Has a conditioning-window baseline visit (age 5-9 y) | 39,301 | 40.5 |
| Has a target-age outcome visit at 12 years | 22,079 | 22.8 |
| Has a target-age outcome visit at 15 years | 14,861 | 15.3 |
| Has a target-age outcome visit at 18 years | 4,731 | 4.9 |
| Final analytic frame (either target age) | 4,857 | 5.0 |
| ***Panel B. Children retained in the final analytic frame, by sex and target age*** |  |  |
| **Sex** | **Target age, y** | **n (children x target age rows)** |
| Male | 12 | 2,445 |
| Male | 15 | 194 |
| Female | 12 | 2,410 |
| Female | 15 | 190 |

**Supplementary Table S2.** City composition of the analytic sample by target age, against each city’s share of the full cohort.

| City | Full cohort, % (n) | Target age 12, % of analytic sample | Target age 15, % of analytic sample | Representativeness ratio, age 12 | Representativeness ratio, age 15 |
| --- | --- | --- | --- | --- | --- |
| Hanoi (HHN) | 76.4 (74,389) | 56.1 | 0.0 | 0.73 | - |
| Ho Chi Minh City (HCP) | 18.7 (18,187) | 34.3 | 89.3 | 1.83 | 4.78 |
| Haiphong (HHP) | 4.9 (4,732) | 9.6 | 10.7 | 1.96 | 2.18 |

The representativeness ratio is each city's percent share of the analytic sample divided by its percent share of the full cohort, a ratio of 1.0 would mean the city is neither over- nor under-represented. Hanoi contributes no children at all to the target-age-15 analytic sample despite being the largest recruitment city in the full cohort (dash indicates the ratio is undefined at zero).

**Supplementary Table S3.** Children included in the final analytic frame versus excluded.

| Characteristic | Excluded (N = 70,057) | Included (N = 4,857) | Total (N = 74,914) | P value |
| --- | --- | --- | --- | --- |
| Sex, n (%) |  |  |  | 0.020 |
| Male | 36,503 (52.1%) | 2,447 (50.4%) | 38,950 (52.0%) |  |
| Female | 33,554 (47.9%) | 2,410 (49.6%) | 35,964 (48.0%) |  |
| City, n (%) |  |  |  | < 0.001 |
| Hanoi (HHN) | 53,299 (76.1%) | 2,716 (55.9%) | 56,015 (74.8%) |  |
| Ho Chi Minh City (HCP) | 13,312 (19.0%) | 1,682 (34.6%) | 14,994 (20.0%) |  |
| Haiphong (HHP) | 3,446 (4.9%) | 459 (9.5%) | 3,905 (5.2%) |  |
| Number of visits, median (Q1-Q3) | 2.0 (1.0, 4.0) | 5.0 (5.0, 7.0) | 2.0 (1.0, 4.0) | < 0.001 |
| Follow-up duration, years, median (Q1-Q3) | 0.9 (0.0, 3.0) | 5.1 (5.0, 6.1) | 1.1 (0.0, 3.1) | < 0.001 |
| First visit year, median (Q1-Q3) | 2020 (2019, 2023) | 2020 (2019, 2020) | 2020 (2019, 2023) | < 0.001 |
| Last visit year, median (Q1-Q3) | 2024 (2020, 2025) | 2025 (2025, 2025) | 2025 (2020, 2025) | < 0.001 |
| Baseline height-for-age Z, median (Q1-Q3) | 0.116 (-0.507, 0.757) | 0.326 (-0.323, 0.999) | 0.142 (-0.493, 0.785) | < 0.001 |
| Baseline BMI-for-age Z, median (Q1-Q3) | 0.242 (-0.587, 1.526) | 0.513 (-0.484, 1.940) | 0.275 (-0.577, 1.582) | < 0.001 |

Every child with at least one merged height-for-age and BMI-for-age visit (N = 74,914) is classified as included in or excluded from the final analytic frame. Baseline height-for-age and BMI-for-age Z are missing for 35,613 excluded children who never had a visit in the age 5 to 9 year conditioning window at all; rows report the characteristics of the remainder. P values are from the Kruskal-Wallis test for continuous variables and the chi-square test for categorical variables, comparing included and excluded children.

**Supplementary Table S4.** Visit density and follow-up duration underlying each target-age submodel.

| Sex | Target age, y | n | Visits, median (IQR) | Follow-up, years, median (IQR) | Baseline-to-target gap, years, median (IQR) | Actual outcome age, years, median |
| --- | --- | --- | --- | --- | --- | --- |
| Male | 12 | 2,445 | 5 (2) | 5.1 (1.1) | 5.0 (0.9) | 11.80 |
| Male | 15 | 194 | 8 (1) | 7.1 (0.0) | 7.1 (0.0) | 14.35 |
| Female | 12 | 2,410 | 5 (2) | 5.1 (1.1) | 5.0 (1.0) | 11.80 |
| Female | 15 | 190 | 8 (1) | 7.1 (0.0) | 7.1 (0.0) | 14.40 |

**Supplementary Table S5.**
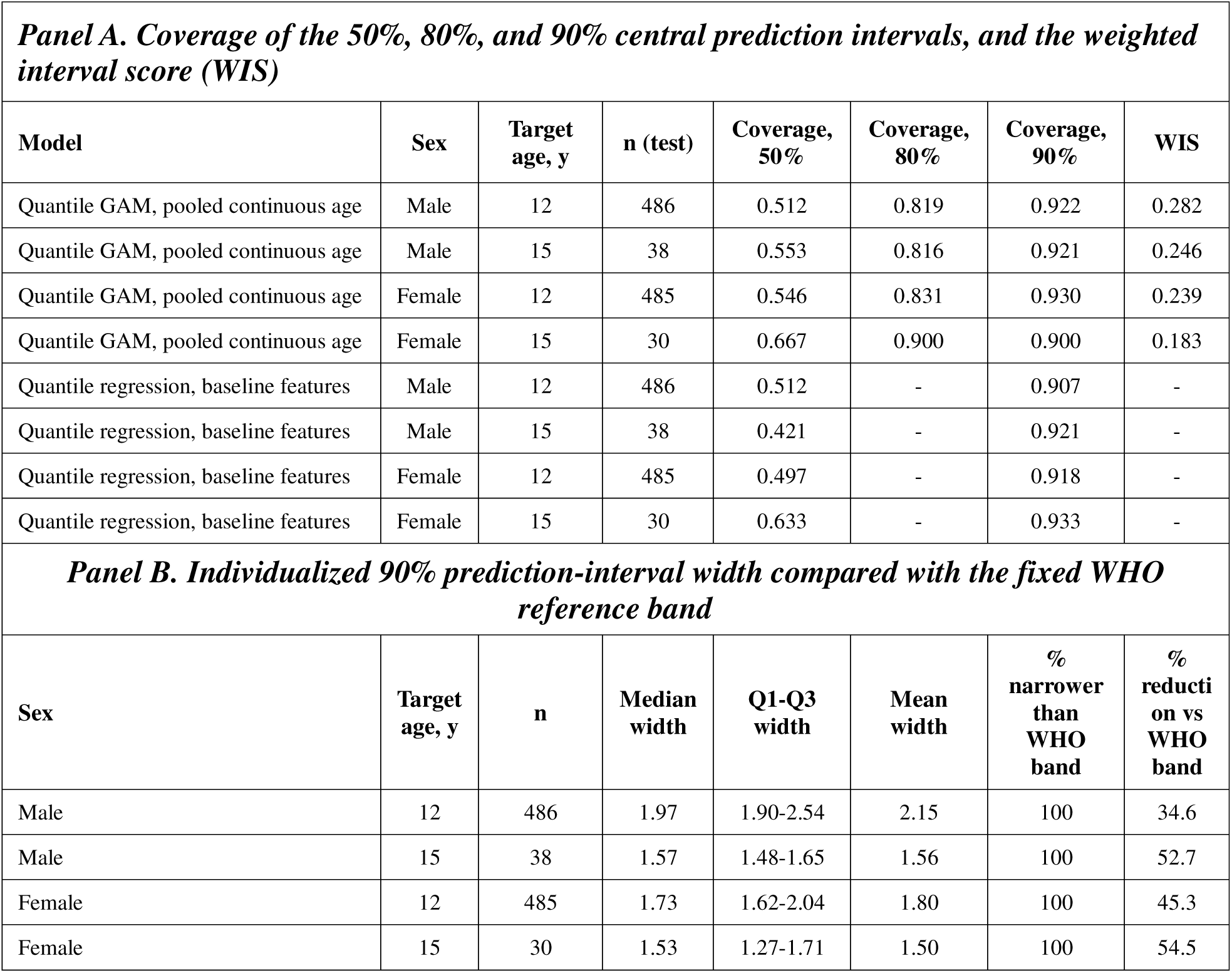
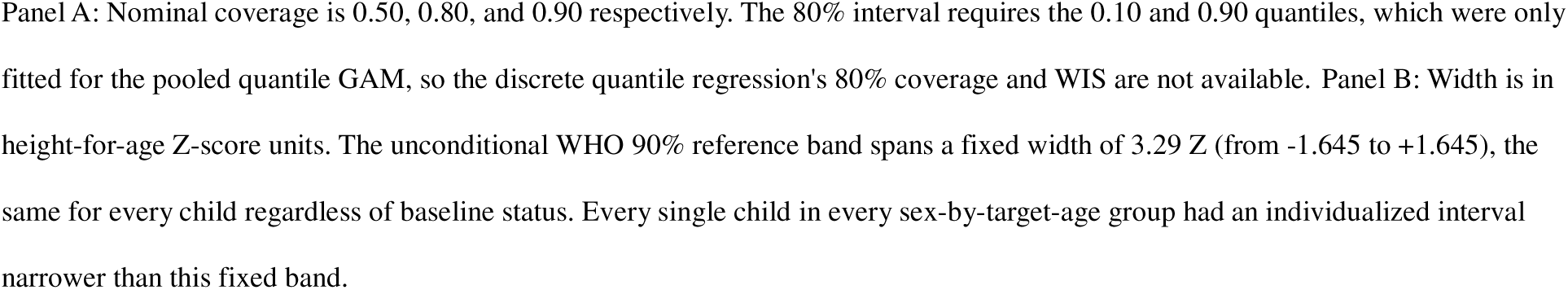
Multi-level interval calibration, weighted interval score, and interval width relative to the WHO reference band.|.

| ***Panel A. Coverage of the 50%, 80%, and 90% central prediction intervals, and the weighted interval score (WIS)*** |  |  |  |  |  |  |  |
| --- | --- | --- | --- | --- | --- | --- | --- |
| Model | Sex | Target age, y | n (test) | Coverage, 50% | Coverage, 80% | Coverage, 90% | WIS |
| Quantile GAM, pooled continuous age | Male | 12 | 486 | 0.512 | 0.819 | 0.922 | 0.282 |
| Quantile GAM, pooled continuous age | Male | 15 | 38 | 0.553 | 0.816 | 0.921 | 0.246 |
| Quantile GAM, pooled continuous age | Female | 12 | 485 | 0.546 | 0.831 | 0.930 | 0.239 |
| Quantile GAM, pooled continuous age | Female | 15 | 30 | 0.667 | 0.900 | 0.900 | 0.183 |
| Quantile regression, baseline features | Male | 12 | 486 | 0.512 | - | 0.907 | - |
| Quantile regression, baseline features | Male | 15 | 38 | 0.421 | - | 0.921 | - |
| Quantile regression, baseline features | Female | 12 | 485 | 0.497 | - | 0.918 | - |
| Quantile regression, baseline features | Female | 15 | 30 | 0.633 | - | 0.933 | - |
| ***Panel B. Individualized 90% prediction-interval width compared with the fixed WHO reference band*** |  |  |  |  |  |  |  |
| Sex | Target age, y | n | Median width | Q1-Q3 width | Mean width | % narrower than WHO band | % reduction vs WHO band |
| Male | 12 | 486 | 1.97 | 1.90-2.54 | 2.15 | 100 | 34.6 |
| Male | 15 | 38 | 1.57 | 1.48-1.65 | 1.56 | 100 | 52.7 |
| Female | 12 | 485 | 1.73 | 1.62-2.04 | 1.80 | 100 | 45.3 |
| Female | 15 | 30 | 1.53 | 1.27-1.71 | 1.50 | 100 | 54.5 |

Panel A: Nominal coverage is 0.50, 0.80, and 0.90 respectively. The 80% interval requires the 0.10 and 0.90 quantiles, which were only fitted for the pooled quantile GAM, so the discrete quantile regression's 80% coverage and WIS are not available. Panel B: Width is in height-for-age Z-score units. The unconditional WHO 90% reference band spans a fixed width of 3.29 Z (from -1.645 to +1.645), the same for every child regardless of baseline status. Every single child in every sex-by-target-age group had an individualized interval narrower than this fixed band.

**Supplementary Table S6.**
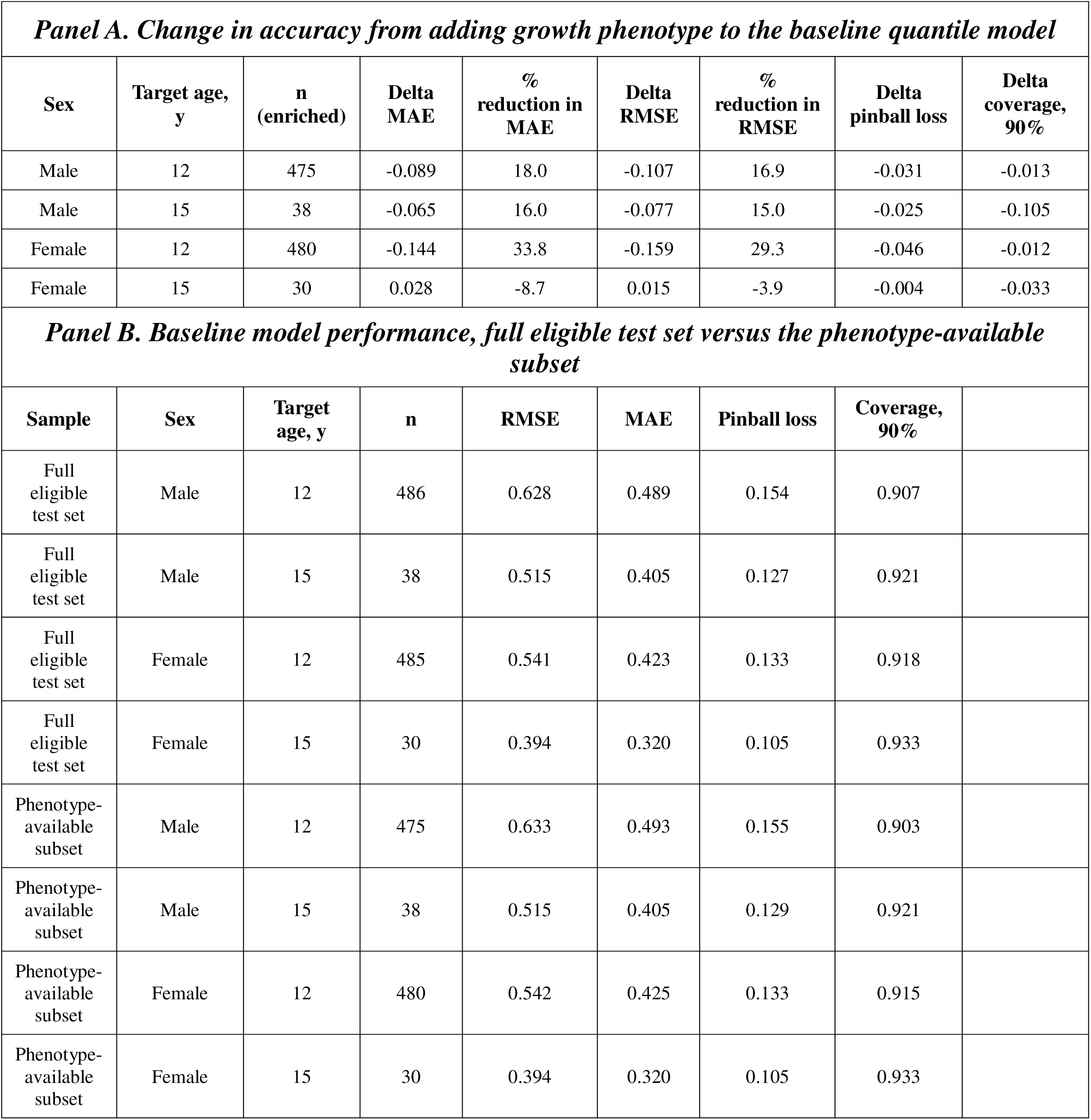
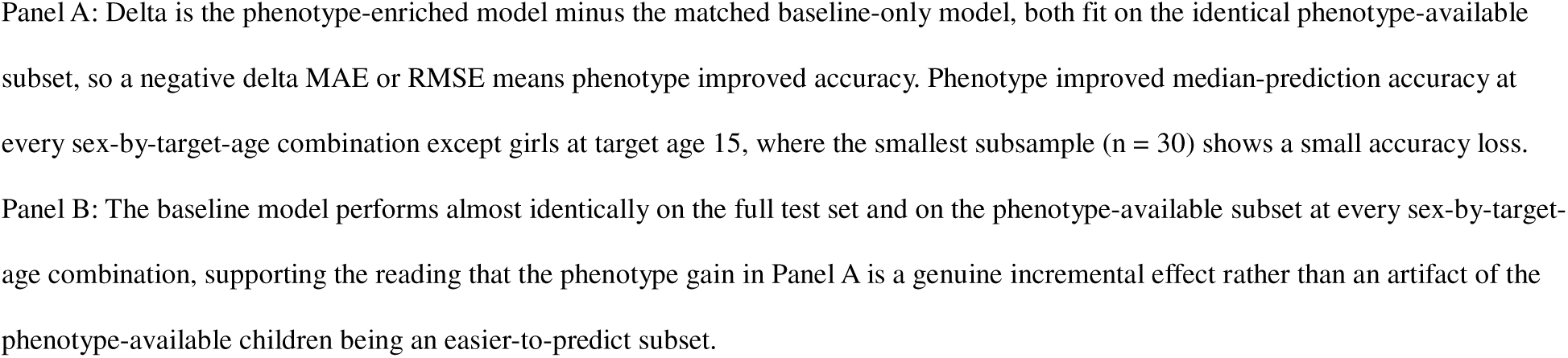
Incremental value of the growth phenotype, and a selection-effect check.

| ***Panel A. Change in accuracy from adding growth phenotype to the baseline quantile model*** |  |  |  |  |  |  |  |  |
| --- | --- | --- | --- | --- | --- | --- | --- | --- |
| **Sex** | **Target age, y** | **n (enriched)** | **Delta MAE** | **% reduction in MAE** | **Delta RMSE** | **% reduction in RMSE** | **Delta pinball loss** | **Delta coverage, 90%** |
| Male | 12 | 475 | -0.089 | 18.0 | -0.107 | 16.9 | -0.031 | -0.013 |
| Male | 15 | 38 | -0.065 | 16.0 | -0.077 | 15.0 | -0.025 | -0.105 |
| Female | 12 | 480 | -0.144 | 33.8 | -0.159 | 29.3 | -0.046 | -0.012 |
| Female | 15 | 30 | 0.028 | -8.7 | 0.015 | -3.9 | -0.004 | -0.033 |
| ***Panel B. Baseline model performance, full eligible test set versus the phenotype-available subset*** |  |  |  |  |  |  |  |  |
| **Sample** | **Sex** | **Target age, y** | **n** | **RMSE** | **MAE** | **Pinball loss** | **Coverage, 90%** |  |
| Full eligible test set | Male | 12 | 486 | 0.628 | 0.489 | 0.154 | 0.907 |  |
| Full eligible test set | Male | 15 | 38 | 0.515 | 0.405 | 0.127 | 0.921 |  |
| Full eligible test set | Female | 12 | 485 | 0.541 | 0.423 | 0.133 | 0.918 |  |
| Full eligible test set | Female | 15 | 30 | 0.394 | 0.320 | 0.105 | 0.933 |  |
| Phenotype-available subset | Male | 12 | 475 | 0.633 | 0.493 | 0.155 | 0.903 |  |
| Phenotype-available subset | Male | 15 | 38 | 0.515 | 0.405 | 0.129 | 0.921 |  |
| Phenotype-available subset | Female | 12 | 480 | 0.542 | 0.425 | 0.133 | 0.915 |  |
| Phenotype-available subset | Female | 15 | 30 | 0.394 | 0.320 | 0.105 | 0.933 |  |

Panel A: Delta is the phenotype-enriched model minus the matched baseline-only model, both fit on the identical phenotype-available subset, so a negative delta MAE or RMSE means phenotype improved accuracy. Phenotype improved median-prediction accuracy at every sex-by-target-age combination except girls at target age 15, where the smallest subsample (n = 30) shows a small accuracy loss.
Panel B: The baseline model performs almost identically on the full test set and on the phenotype-available subset at every sex-by-target-age combination, supporting the reading that the phenotype gain in Panel A is a genuine incremental effect rather than an artifact of the phenotype-available children being an easier-to-predict subset.

**Supplementary Table S7.**
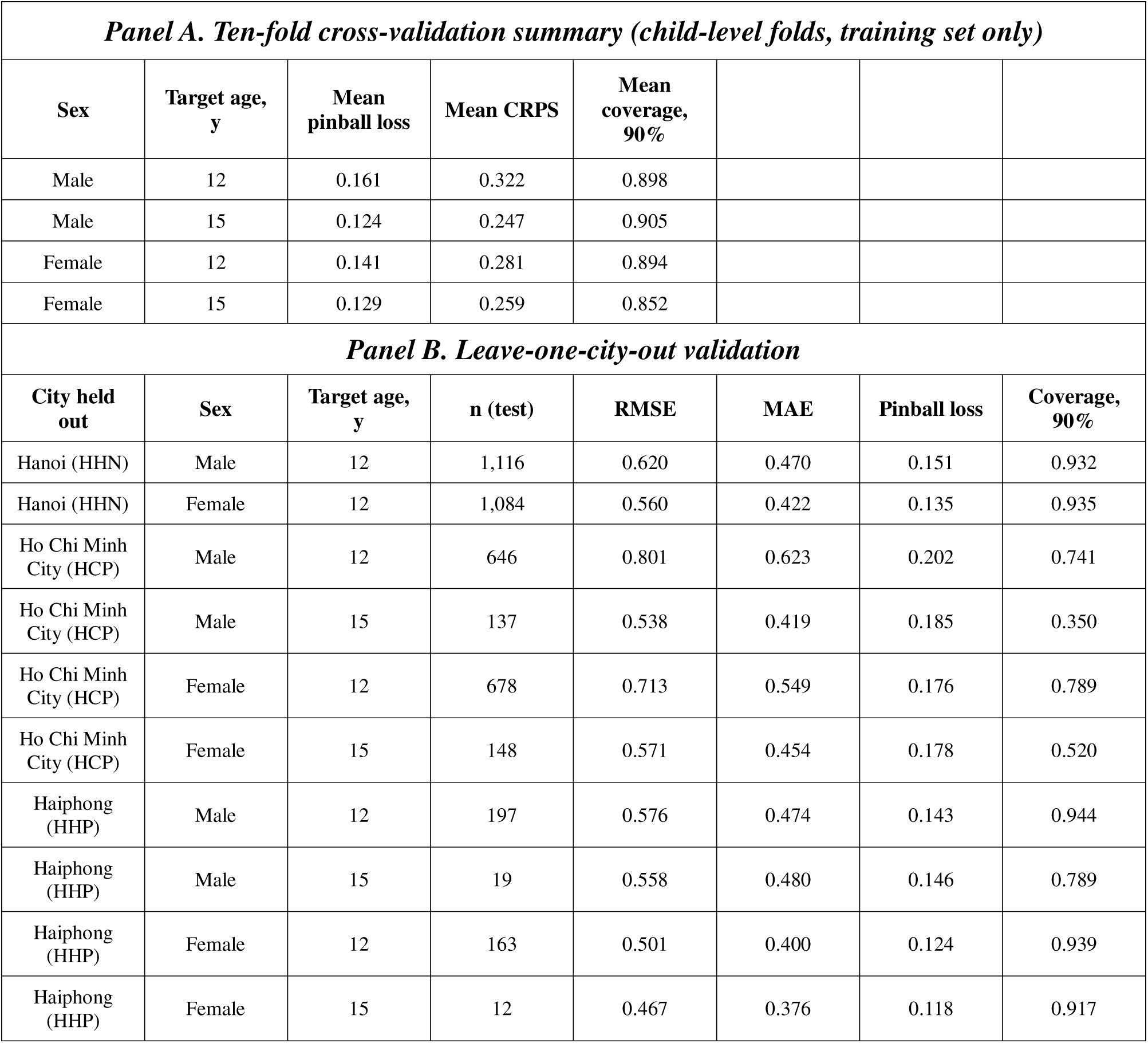
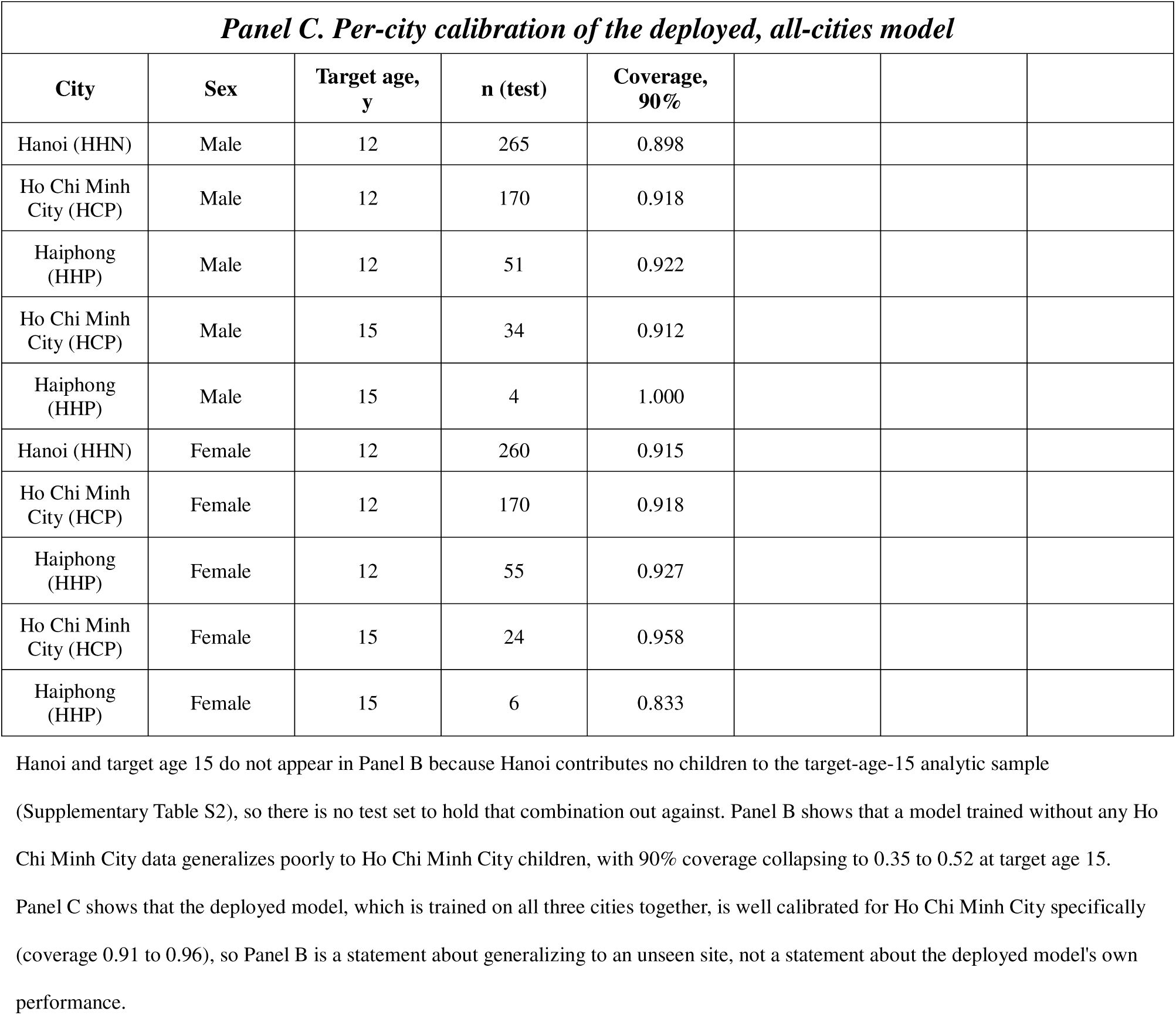
Cross-validation, leave-one-city-out validation, and per-city calibration of the deployed model.

**Supplementary Table S8.** Worked clinical examples.

| **Case** | **Baseline HAZ** | **Baseline BAZ** | **Target age, y** | **Predicted 5th percentile** | **Predicted median** | **Predicted 95th percentile** |
| --- | --- | --- | --- | --- | --- | --- |
| Boy, average height, mild overweight at age 6, HCP site | 0.0 | 0.3 | 12 | -1.07 | 0.08 | 1.49 |
| Boy, average height, mild overweight at age 6, HCP site | 0.0 | 0.3 | 15 | -1.11 | 0.10 | 0.63 |
| Boy, short for age, high BMI at age 6, HCP site | -1.5 | 1.8 | 12 | -2.07 | -1.09 | 0.33 |
| Boy, short for age, high BMI at age 6, HCP site | -1.5 | 1.8 | 15 | -2.57 | -1.10 | -0.92 |
| Boy, tall for age, normal BMI at age 6, HCP site | 1.5 | 0.2 | 12 | 0.03 | 1.32 | 2.70 |
| Boy, tall for age, normal BMI at age 6, HCP site | 1.5 | 0.2 | 15 | 0.28 | 1.28 | 2.03 |

All three cases are predicted from the primary discrete quantile regression model, holding city at Ho Chi Minh City. The short-for-age, high-BMI case is the only one whose entire predicted 5th to 95th percentile interval remains below zero by target age 15, illustrating that the model predicts sustained short stature with high confidence for this specific combination of baseline height and BMI status, a distinction a single fixed WHO percentile chart cannot make since it does not condition on BMI at all.

